# Evaluating 5-Item Short Forms of Youth Depression Measures

**DOI:** 10.64898/2026.10.13.26365293

**Authors:** Allison M. Loomis, Phoebe A. Rodda, Jennifer K. Youngstrom, Robert L. Findling, A. John Rush, Eric A. Youngstrom

## Abstract

Brief assessments reduce clinician and patient burden and increase utilization. Recently, five core depression symptoms (low mood, poor concentration, low self-worth, loss of interest, low energy) have been shown to be particularly sensitive to treatment effects in adults. How these core depressive symptom domains perform for screening or diagnostic purposes in youth is unknown. To address this question, we extracted five analogous symptom items from four widely used symptom assessment tools for children and adolescents (Child Behavior Checklist [CBCL], Youth Self Report [YSR], Parent General Behavior Inventory [P-GBI], and Adolescent General Behavior Inventory [A-GBI]) to create 5-item short forms. We evaluated their psychometric properties and diagnostic utility in a large (*N*=1185), racially diverse sample (48% Black youth) across community and academic outpatient mental health settings, with diagnoses established through LEAD consensus. All short forms correlated highly with full-length measures (*r*=.74-.92) and demonstrated comparable inter-item correlations (*r* =.19-.46). The short forms did not differ meaningfully from the full measures in correlations with psychiatric diagnoses, demographics, or diagnostic interviews. Area under the curve (AUC) values for identifying depression and any mood disorder diagnosis (determined by semi-structured diagnostic interviews) were fair to good (AUC=0.60-0.79), which were not worse than full measures, except for the P-GBI. The CBCL-5, YSR-5, P-GBI-5, and A-GBI-5 may be useful brief depression assessments in youth. How these five symptom domains perform as outcome measures in treatment deserves study.

## Evaluating 5-Item Short Forms of Youth Depression Measures

Rates of mood disorders have increased 1.5- to 2-fold over the past decade (Lu, 2019; Thapar et al., 2022), and up to 10% of children and adolescents meet criteria for a mood disorder in a given year (Kessler et al., 2012). Major depressive disorder (MDD) is particularly prevalent, with an estimated lifetime prevalence of 19% among adolescents (Shorey et al., 2022). Mood disorders are associated with functional impairment and suicide risk (Nagar et al., 2010), underscoring the importance of early assessment, accurate identification, and treatment.

Systematic assessment informs treatment for depression in two main ways. First, youth and their caregivers complete assessments to screen for and evaluate mood disorder symptoms, helping determine whether treatment or intervention is needed. Second, assessments administered frequently throughout treatment can evaluate progress and inform treatment decisions (i.e., measurement-based care [MBC], Trivedi et al., 2006). Routinely assessing symptoms leads to quicker alleviation of symptoms than impressionistic clinical judgments for depressed patients (Guo et al., 2015; Husain et al., 2025). While evidence indicates the value of implementing systematic assessments, their administration can impose a time burden on patients, their caregivers, and clinicians. Further, while longer assessments are more likely to capture broad symptom domains, they are also less likely to be used (Rolstad et al., 2011). Thus, balancing coverage and diagnostic/progress accuracy with brevity is pivotal.

The Very Quick Inventory of Depressive Symptomatology (VQIDS-5; De La Garza et al., 2017) aims to balance these domains by assessing only five core symptoms of depression. The original five were (1) low mood, (2) psychomotor retardation, (3) low self-worth, (4) loss of interest, and (5) low energy (Bech et al., 1975). These symptoms, plus an anxiety item, were the only Hamilton Rating Scale for Depression (HRSD; Hamilton, 1960) items that met criteria for calibration, ascending monotonicity, and dispersion. Since then, two important changes have been made to the VQIDS-5. First, the somatic anxiety item identified by Bech and colleagues was removed because it is not a criterion symptom of depression and had the lowest factor loading among the HRSD_6_ items in principal component analysis (Bech et al., 2011). Second, “lack of concentration” replaced “psychomotor retardation” because concentration was more sensitive to treatment effects (Rush et al., 2026). Therefore, the five core symptoms of depression, supported by a body of evidence, are (1) low mood, (2) difficulties in concentration, (3) low self-worth, (4) loss of interest, and (5) low energy. Both self-report (VQIDS-SR_5_) and clinician-report (VQIDS-C_5_) versions of the scales focused on these five symptoms have shown sensitivity to change comparable to the self- and clinician-report versions of the Quick Inventory of Depressive Symptomatology (QIDS-SR_16_ and QIDS-C_16_; Rush et al., 2003) in both adults and youth aged 8 to 17 years (Croarkin et al., 2025; Rush et al., 2022). Additionally, VQIDS-SR_5_ scores have demonstrated strong item-total correlations, high correlations with QIDS-SR_16_ scores, and reliable conversion to Patient Health Questionnaire (PHQ-9; Kroenke et al., 2001) scores, with similar sensitivity to depressive symptoms in adolescents (Nandy et al., 2024). At less than a third of the length (5 versus 16 items), the VQIDS-5 may reduce the time burden of measurement while still producing valid and reliable scores.

Although the VQIDS-5 provides a quick, efficient, and reliable tool for monitoring changes in core depressive symptoms during treatment, longer measures remain commonly used for symptom inventory, diagnosis, and screening. A few of the most comprehensive and systematically validated assessment tools for mood disorders in youth include the Achenbach System of Empirically Based Assessment (ASEBA; Achenbach and Rescorla, 2001) and the General Behavior Inventory (GBI; Depue et al., 1981). These tools capture a broad range of behavioral and emotional challenges but may be too lengthy for frequent progress monitoring. Since these measures all include symptoms that match the core depressive features, it may be possible to focus exclusively on these five symptoms to obtain quick screening information to inform diagnosis and treatment planning, or to extract a short form for treatment follow-up.

Previous short forms have been developed, though none are as brief or guided by the framework described above. Achenbach and Rescorla’s (2011) 19-item Brief Problem Monitor includes six internalizing items but assesses internalizing psychopathology broadly rather than depression specifically. Three 10-item GBI short forms have also been developed: one assessing mania (P-GBI10M; Youngstrom et al., 2008) and two assessing depression (Youngstrom et al., 2020, 2021). Items were selected to optimize discrimination between depressive or bipolar disorders and other psychiatric conditions while balancing internal consistency, content coverage, and criterion validity (Youngstrom et al., 2020, 2021). Seven-item mania and depression scales were similarly designed to maximize internal consistency and can be administered separately or together to preserve the GBI’s original factor structure (Youngstrom et al., 2020, 2021). Although substantially shorter than the 28-item hypomanic/biphasic and 46-item depression scales, these forms remain longer than the five-item measures examined here.

It is possible that five items selected to map onto core depression symptoms could offer similar reliability, content coverage, and discriminative validity while reducing the length of measures. Using core symptoms assessed across commonly used depression measures could facilitate cross-scale linking and sample calibration (Bech et al., 2011; De La Garza et al., 2017; Fried, 2017). There is low item content overlap among different depression measures that are often used interchangeably, raising important concerns about the replication of results (Fried, 2017). Selecting items for new short forms based solely on EFA may exacerbate this low content overlap, leading to sample-specific results that may not replicate. Creating short measures based on the same core symptoms will reduce content discrepancy and ensure that depression measures are rooted in research and theory. Given the growing empirical support for core MDD symptoms, using these items to create short forms is likely to increase the generalizability of results across samples (De La Garza et al., 2017). Further, using similar items across depression scales facilitates the translation and comparison of scores in clinical settings, making evaluation and progress monitoring more efficient and accurate.

The utility of the five core symptoms for diagnosis and risk assessment remains unclear, as the VQIDS-5 was originally designed as a brief outcome measure and has not been established as a diagnostic tool (De La Garza et al., 2017). In contrast, the CBCL, YSR, and GBI have been used as symptom inventories and for diagnostic assessment. The current study tested whether the five VQIDS-5 symptoms could support diagnosis and risk assessment, potentially providing a brief alternative to longer symptom inventories. We developed the five-item short forms from the CBCL, YSR, P-GBI, and A-GBI by matching items to the five core symptoms and selecting the strongest-loading item when multiple items assessed the same symptom. We compared short and full measures on reliability, factor loadings, criterion validity, and diagnostic accuracy, and examined potential scoring thresholds using kappa coefficients and likelihood ratios. To our knowledge, this is the first study to construct 5-item short forms from preexisting diagnostic and risk assessment measures in youths. Further, we seek to build upon existing VQIDS-5 literature by examining the ability of similar 5-item short forms to assess risk of mood disorder diagnosis generally, in addition to unipolar depression.

## Methods

### Participants

We conducted secondary analyses of families presenting to either a community mental health center or an academic medical center in the same Midwestern city. Youth aged 4 to 18 (Academic: *M_age_*=11.53, *SD_age_*_=_3.37; Community: *M_age_*=10.89, *SD_age_*=3.20) were recruited, along with their caregivers, between 1998 and 2008. However, only youth 11 years and older completed self-report measures. Youth completed the YSR as part of a clinical activity, so although the research protocol was intended for ages 11 and older, four younger cases (ages 9-10) were available for secondary analysis because the intake clinician chose to collect the information. *N*=1185 caregivers, including 628 from the academic medical center and 557 from the community, completed the five short-form items on both the CBCL and P-GBI and were thus included in all parent-report analyses. 560 youth, including 251 from the academic medical center and 309 from the community, completed the five items on both the YSR and A-GBI and were thus included in all youth-report analyses. In the academic sample, 60.67% (*n*=381) were male, and in the community sample, 59.25% were male. Regarding race in the academic sample, 79% (*n*=495) of participants were White, 13% (*n*=79) were Black, and 8% (*n*=54) identified as Native American, Alaskan Native, Asian, Pacific Islander, Hispanic, or “Other”. In the community sample, 89% (*n*=495) of participants were Black, 6% (*n=*36) were White, and 4% (*n*=26) identified as Native American, Alaskan Native, Asian, Pacific Islander, Hispanic, or “Other.”

Exclusion criteria included (a) a diagnosis or documented history of pervasive developmental disorder, (b) potential cognitive disability, as indicated by educational history, standardized cognitive ability test scores below 70, or Peabody Picture Vocabulary Test-Third Edition (Dunn and Dunn, 1997) scores below 70, or inability to complete the interview in English.

### Diagnoses

All participants completed a semi-structured diagnostic interview, the Kiddie Schedule for Affective Disorders and Schizophrenia (Geller et al., 2001; Kaufman et al., 1997), administered by a highly trained rater and following the Diagnostic and Statistical Manual of Mental Disorders (4^th^ ed.; DSM-IV; American Psychiatric Association, 1994) diagnostic criteria. All diagnoses were finalized through a LEAD consensus conference (Spitzer, 1983) supervised by a licensed clinical psychologist or a board-certified child psychiatrist.

### Measures

#### Parent and Adolescent General Behavior Inventory (P-GBI; A-GBI)

The GBI (Depue et al., 1981) is a comprehensive, well-validated, and widely used measure of depressive and hypomanic symptoms, with 73 items, 46 of which assess depressive symptoms. These 4-point Likert-type items range from 0 (“never or hardly ever”) to 3 (“very often or almost constantly”), with higher scores indicating greater symptom severity (Depue et al., 1981). An example item is “Have people said that you looked sad or lonely?” The P-GBI (Youngstrom et al., 2001) and A-GBI (Danielson et al., 2003) have nearly identical item content and the same subscale structure as the GBI. The most common modification to the parent version was changing “you” to “your child.” Psychometric analyses support the reliability of the P-GBI and A-GBI in assessing mood disorders in youth, with both scales demonstrating good diagnostic accuracy and the ability to differentiate among unipolar depression, bipolar disorders, disruptive behavior disorder, ADHD, and no diagnosis (Alcaino et al., 2024; Danielson et al., 2003; Youngstrom et al., 2015a, 2001).

#### Child Behavior Checklist (CBCL) and CBCL for Youth Self Report (YSR)

The CBCL and YSR are part of the Achenbach System of Empirically Based Assessment (Achenbach and Rescorla, 2001) and have been thoroughly validated, with a long history of research and use. The CBCL and YSR each consist of 118 items that ask caregivers or youth to rate symptoms on a scale from 0 (“not true [as far as you know]”) to 2 (“very true or often true”). An example item is “Unhappy, sad, or depressed.” Each instrument comprises eight syndrome scales. The present analysis used the three syndrome scales that comprise the “internalizing” domain: Anxious/Depressed, Withdrawn/Depressed, and Somatic Complaints. Given the participants’ age range, the CBCL for children ages 6 to 18 was used. The YSR was administered only to youth aged 11 and older, as use in children aged 10 and younger is not recommended due to reading-level concerns. The CBCL and YSR demonstrate good reliability and validity and have empirical support for their diagnostic validity and utility in measuring both internalizing and externalizing domains in youth (Achenbach and Rescorla, 2007; Cohen et al., 1985).

#### Mood Severity Ratings

Two mood severity measures were used to establish criterion validity of the short forms. The Young Mania Rating Scale (YMRS; Young et al., 1978) is a semi-structured interview consisting of 11 items. Items are rated on either a 0-to-4 or a 0-to-8 scale and combine youth and caregiver reports of mood symptoms with clinician observations. Second, the Child Depression Rating Scale-Revised (CDRS-R; Poznanski et al., 1984) is a semi-structured interview consisting of 17 items. Items are rated on either a 1-to-5 or a 1-to-7 scale and combine youth and caregiver reports with clinician observations.

### Procedure and Data Analytic Plan

#### Choosing Items on the Short Forms

Five core depression symptoms were extracted from the VQIDS-5: (1) sad mood, (2) negative self-outlook, (3) lack of involvement/interest, (4) fatigue/low energy, and (5) lack of concentration. We evaluated all items on the CBCL, YSR, P-GBI, and A-GBI to compile a list of items (“contender list”) corresponding to each of the five core depression symptoms. For the CBCL and YSR, we observed a clear one-to-one match, with only five items mapping onto the five core depression symptoms. These five items were present in both the CBCL and YSR and were used to create short versions of these measures (CBCL-5; YSR-5). Matching symptoms to the P-GBI and A-GBI was more complex because four of the five symptoms (all except lack of involvement/interest) mapped onto multiple P-GBI or A-GBI items. To choose among similar items, we performed an exploratory factor analysis (EFA) on all items from the contender list across all symptoms (see Supplementary Table S1). Average scores for the short forms were computed using pro-rated scores if participants completed four of the five items, consistent with clinical convention.

#### Psychometric Properties

We assessed correspondence between the short and full measures using Pearson correlations and compared them with projected correlations for five randomly selected items. We evaluated internal consistency using Cronbach’s alpha and compared observed values with projected alpha values for randomly selected five-item subsets. Because alpha is sensitive to scale length, we also calculated mean inter-item correlations (IICs) for the short and full measures. Finally, we examined the short forms’ factor loadings using principal-axis factor analysis with Promax rotation.

#### Criterion Validity

We evaluated criterion validity by correlating the short and full measures with psychiatric diagnoses, demographic characteristics, and YMRS and CDRS-R total scores. We then quantified differences between corresponding correlations using Cohen’s *q*. Within each criterion domain—clinical severity, diagnosis, and demographics—we ordered correlations by expected strength of association with short-form scores, following Youngstrom et al. (2020). To account for multiple comparisons, we applied a setwise Bonferroni correction within each of the four short forms across the 14 criteria. Cohen’s *q* values below .10 were considered small, indicating no clinically meaningful difference between the short-and full-form correlations (Cohen, 1988).

#### Diagnostic Prediction Accuracy

To evaluate the ability of the short forms to predict any mood disorder or MDD diagnosis accurately, and to compare their performance with their corresponding full-length measures, we calculated area under the curve (AUC) and Cohen’s *d* values for both the 5-item and full-length measures’ prediction of (1) any mood disorder diagnosis and (2) MDD diagnosis with the academic and community samples, separately. “Any mood disorder” refers to youth who met DSM-IV (American Psychiatric Association, 1994) diagnostic criteria for depression, dysthymic/persistent depressive disorder, otherwise specified depressive disorder, or bipolar disorder. Finally, for each comparison between a short and a full-length measure, we reported DeLong’s test to estimate the effect size of the difference in AUC between them.

#### Likelihood Ratio Tests

Lastly, we calculated multilevel likelihood ratios for the short forms in detecting (1) any mood disorder diagnosis and (2) an MDD diagnosis using the overall sample. These ratios are based on the sensitivity and specificity of the short forms across different score ranges. For each score range, we calculated the likelihood ratio using the lowest score in that range as the cut point.

## Results

### Psychometric Properties

Table 1 summarizes descriptive statistics and the psychometric properties of the short forms, including projected and observed correlations with full-length measures, average inter-item correlations, projected and observed internal consistency, and factor loadings. The CBCL and YSR short forms reduced length by 84%, while the P-GBI-5 and A-GBI-5 reduced length by 89%. The short forms correlated strongly with the full-length measures for the CBCL (academic *r=*.74; community *r=*.76), YSR (academic *r=*0.84; community *r=*0.82), P-GBI (academic *r=*.90; community *r=*.85), and A-GBI (academic *r=*0.92; community *r*=0.89). These correlations were all higher than the projected correlations between the short and full-length measures.

**Table 1.**
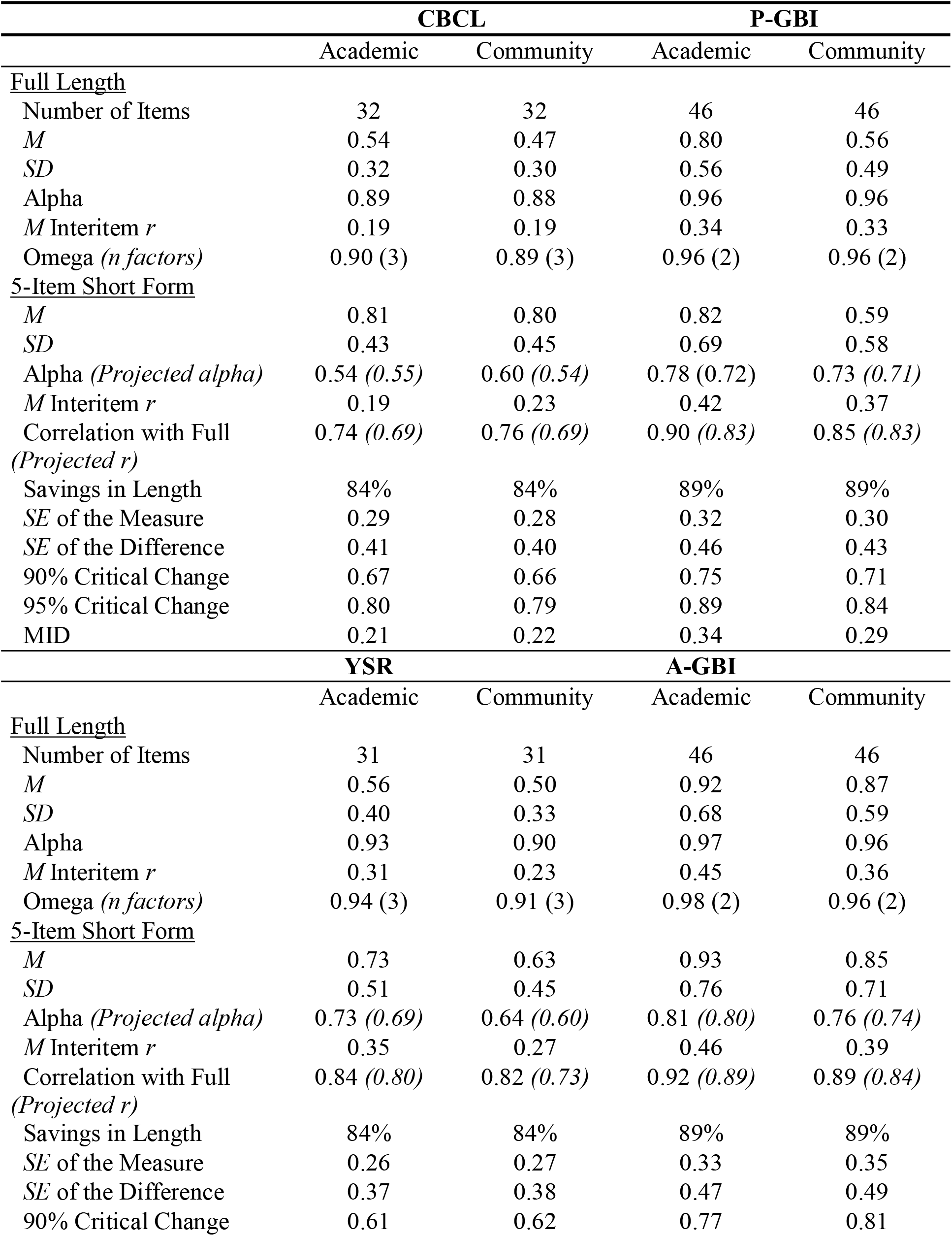
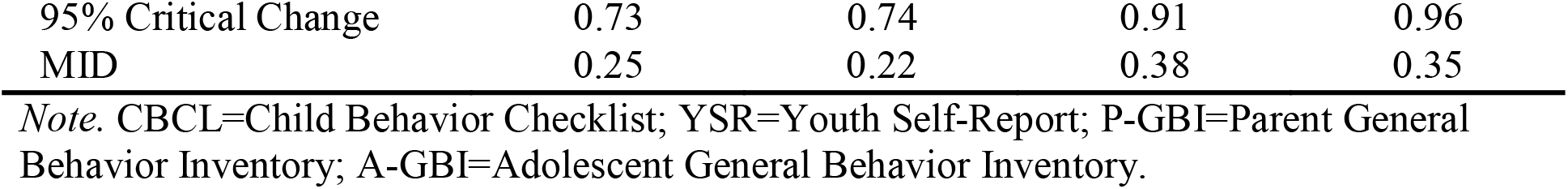
Descriptives and Psychometric Properties.

Additionally, all short forms had average inter-item correlations approximately equal to or higher than those of the full-length measures. The observed internal consistencies of the short forms were generally higher than the projected internal consistencies, except for the CBCL-5 in the academic sample (α_o_=.54, α_p_=.55). Critical values for 90% and 95% confidence that a change in the short forms’ mean scores is likely to reflect a true change in depression symptoms are also reported in Table 1.

Factor loadings are summarized in Table 2. Across scales, 0.33 was used as the minimum threshold for a sufficient factor loading, and loadings above 0.70 were considered strong (Tabachnick and Fidell, 2018). Item 2 on the CBCL-5 (“Can’t concentrate, can’t pay attention for long”) had a poor factor loading of .15 in the academic sample and .07 in the community sample. Item 4 on the CBCL-5 (“There is very little he/she enjoys”) showed a poor factor loading (.17) in the academic sample but was moderately strong in the community sample (.50).

**Table 2.**
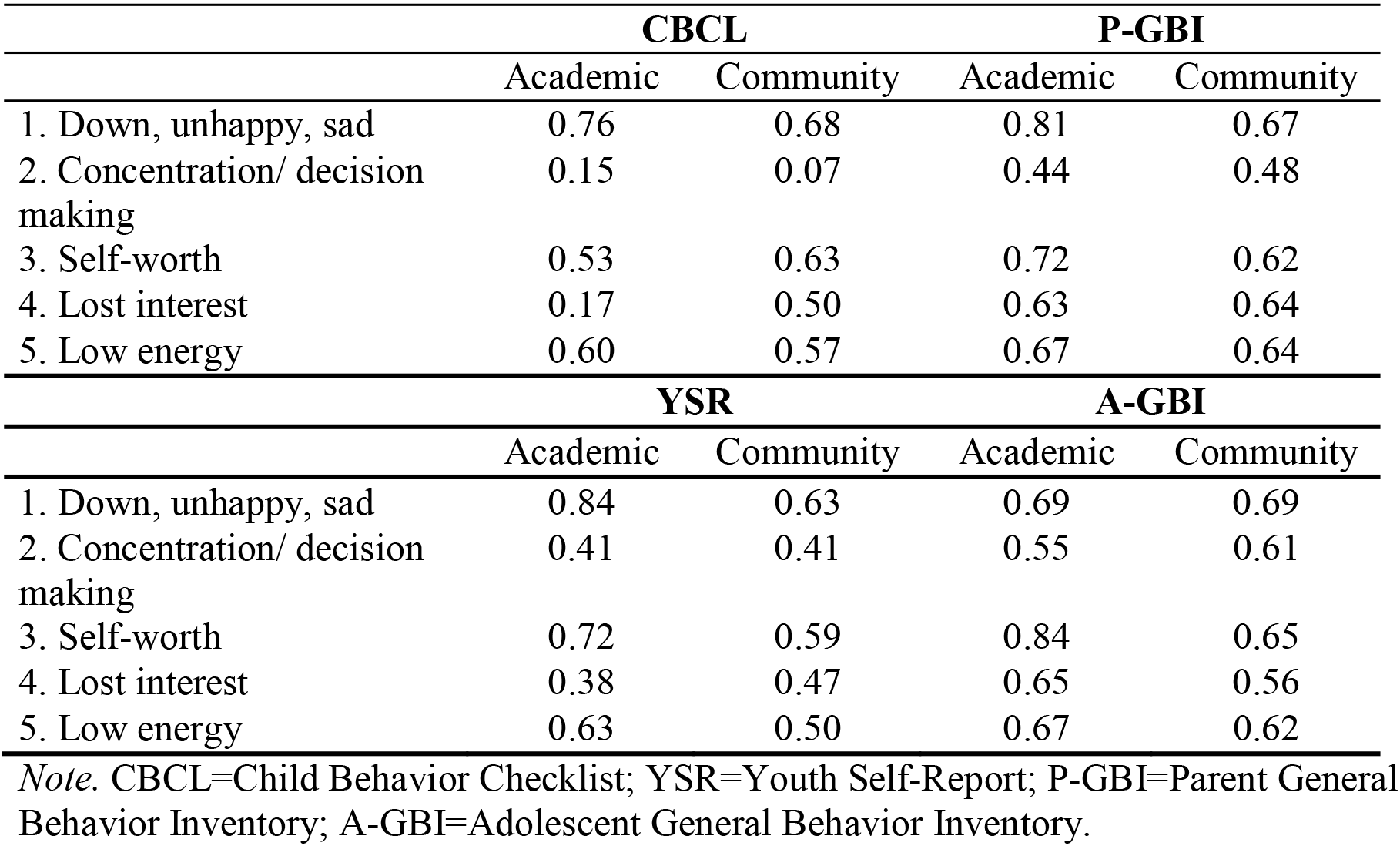
Factor Loadings from Principal Axis Factor Analysis.

The remaining items on the CBCL-5 had moderate-to-strong factor loadings (.53-.76) across samples. All items on the YSR-5, P-GBI-5, and A-GBI-5 had moderate-to-strong factor loadings across samples (.38-84, .44-.81, and .55-.84, respectively).

### Criterion Validity

To evaluate criterion validity of the short forms, we calculated Cohen’s *q* for the difference between correlations for the 5-item and full-length measures across various criteria. Across diagnoses (including ADHD, anxiety disorders, bipolar disorder, conduct disorder, any mood disorder, ODD, PTSD, and unipolar depression), demographics (including sex, age, and race), the YMRS, and the CDRS-R, all *q*s were less than .15 for all 5-item short forms, with most corresponding to small effect sizes less than .10 (Cohen, 1988; see Table 3). This result suggests that there are little-to-no practical differences between the full-length and short-form measures in how they relate to clinical diagnoses, demographics, and diagnostic interviews of depression and other mood disorders.

**Table 3.** Criterion Validity.

| Criterion Variable | 5-item | Full | Cohen's *q* | 5-item | Full | Cohen's *q* |
| --- | --- | --- | --- | --- | --- | --- |
| CBCL |  |  | P-GBI |  |  |  |
| CDRS-R | .43 | .49 | -0.08* | .50 | .58 | -0.11** |
| YMRS | .18 | .21 | -0.03 | .24 | .32 | -0.09** |
| Count of Diagnoses | .26 | .22 | 0.04 | .17 | .21 | -0.04* |
| **Diagnoses** |  |  |  |  |  |  |
| Any Unipolar Depressive Disorder | .25 | .25 | 0.00 | .24 | .27 | -0.03 |
| Any Mood Disorder | .37 | .42 | -0.06 | .43 | .53 | -0.13** |
| Any Bipolar Spectrum | .16 | .22 | -0.06 | .25 | .33 | -0.09** |
| Any Anxiety Disorder | .10 | .18 | -0.08** | .06 | .11 | -0.05* |
| Any ADHD | .08 | -.06 | 0.14** | -.02 | -.08 | 0.06** |
| Any Oppositional Defiant Disorder | .08 | .01 | 0.07* | -.02 | .00 | -0.02 |
| Any Conduct Disorder | .03 | .04 | -0.01 | .02 | .03 | -0.01 |
| Any Posttraumatic Stress Disorder | .07 | .10 | -0.03 | .06 | .09 | -0.03 |
| **Demographics** |  |  |  |  |  |  |
| Age (years) | .17 | .14 | 0.03 | .20 | .21 | -0.01 |
| Female | .07 | .15 | -0.08* | .10 | .14 | -0.04 |
| White | .00 | .01 | -0.01 | .00 | .00 | 0.00 |
| YSR |  |  | A-GBI |  |  |  |
| CDRS-R | .40 | .38 | 0.02 | .37 | .40 | -0.04 |
| YMRS | .13 | .11 | 0.02 | .09 | .10 | -0.01 |
| Count of Diagnoses | .17 | .17 | 0.00 | .13 | .14 | -0.01 |
| **Diagnoses** |  |  |  |  |  |  |
| Any Unipolar Depressive Disorder | .19 | .16 | 0.03 | .16 | .17 | -0.01 |
| Any Mood Disorder | .29 | .25 | 0.04 | .22 | .24 | -0.02 |
| Any Bipolar Spectrum | .12 | .11 | 0.01 | .08 | .09 | -0.01 |
| Any Anxiety Disorder | .17 | .23 | -0.06 | .19 | .20 | -0.01 |
| Any ADHD | -.05 | -.09 | 0.04 | -.11 | -.12 | 0.01 |
| Any Oppositional Defiant Disorder | -.02 | -.04 | 0.02 | -.06 | -.05 | -0.01 |
| Any Conduct Disorder | -.01 | .03 | -0.04 | .03 | .05 | -0.02 |
| Any Posttraumatic Stress Disorder | .14 | .17 | -0.03 | .15 | .15 | 0.00 |
| **Demographics** |  |  |  |  |  |  |
| Age (years) | .13 | .09 | 0.04 | .13 | .15 | -0.02 |
| Female | .30 | .30 | 0.00 | .18 | .27 | -0.09** |
| White | .05 | .01 | 0.04 | .08 | .08 | 0.00 |

**Note.** A Bonferroni correction for the correlations computed within each scale (i.e., CBCL, P-GBI, YSR, and A-GBI) was applied. CBCL=Child Behavior Checklist; YSR=Youth Self-Report; P-GBI=Parent General Behavior Inventory; A-GBI=Adolescent General Behavior Inventory; CDRS-R=Child Depression Rating Scale-Revised; YMRS=Youth Mania Rating Scale; ADHD=attention deficit/hyperactivity disorder.
\* $p < .0036$ , \*\* $p < .00036$ adjusted threshold, two-tailed.

### Diagnosis Prediction Accuracy

Table 4 summarizes the AUC and Cohen’s *d* values for all 5-item and full-length measures in predicting any mood disorder and MDD diagnoses. Guidelines for AUC interpretation were derived from the work of Youngstrom and colleagues (2019) and Rice and Harris (2005). For the 5-item measures, AUC values were generally higher for caregiver-report measures and for predicting any mood disorder diagnosis (rather than predicting a unipolar depression diagnosis specifically). AUC values were mostly good (i.e., between .70 and .79) or excellent (i.e., between .80 and .89) for the CBCL-5 and P-GBI-5, and fair (i.e., between .60 and .69) for the YSR-5 and A-GBI-5 when predicting any mood disorder diagnosis in both the academic and community samples. When predicting a depression diagnosis, short-form performance was mostly fair across samples. There were no significant differences in predictive accuracy for either any mood disorder or a depression diagnosis between the short and full-length versions of the youth-report measures (i.e., YSR and A-GBI) after stratifying by sample population source (i.e., academic versus community). For the P-GBI, predictive ability for any mood disorder diagnosis differed significantly between the short form (academic AUC=0.80, community AUC=0.71) and the full-length measure (academic AUC=0.85, community AUC=0.79). However, despite these significant differences, the P-GBI-5 still demonstrated good predictive ability for any mood disorder diagnosis across samples. Overall, these results support the predictive ability of the 5-item short forms, particularly for any mood disorder diagnosis.

**Table 4.**
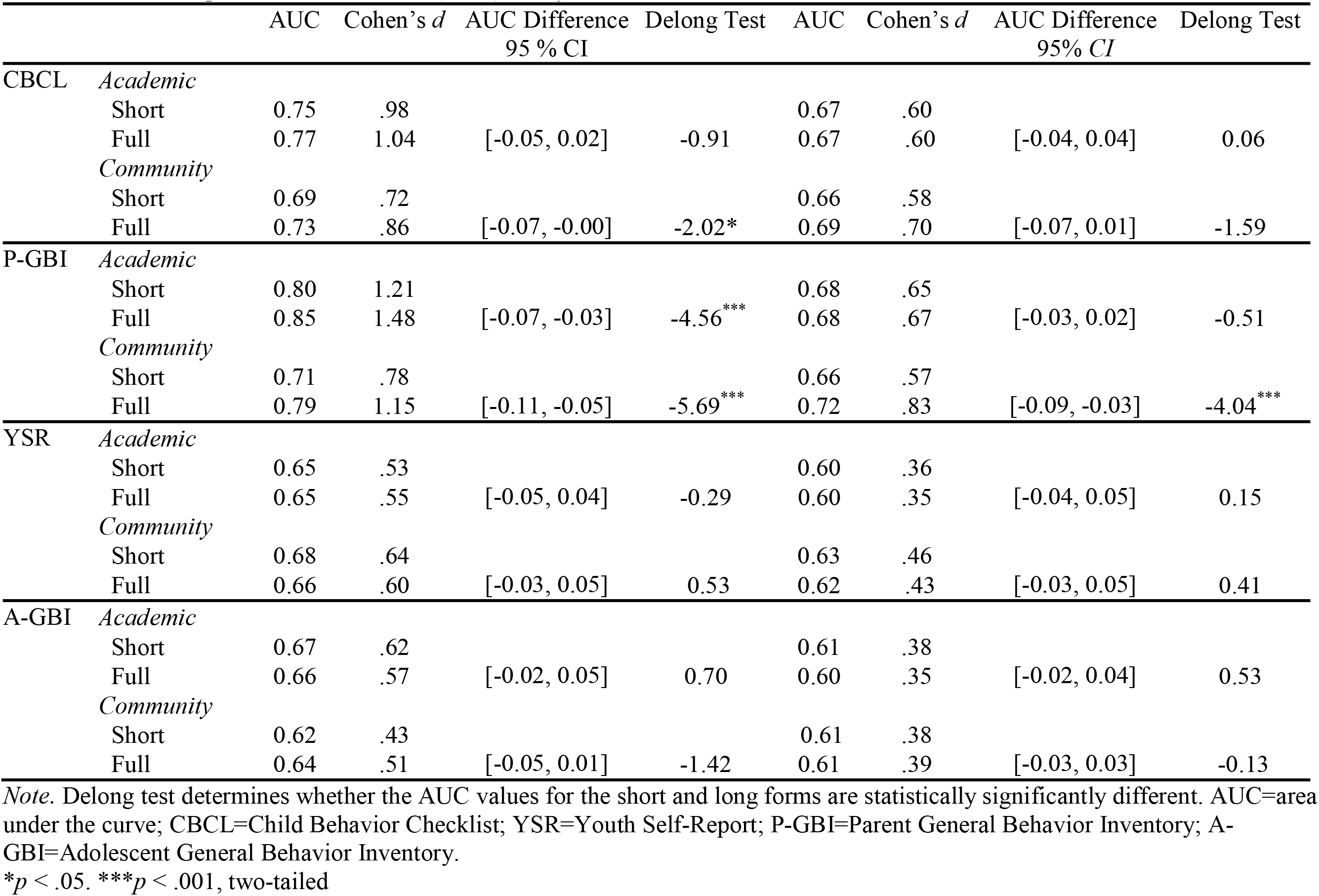
Receiver Operator Characteristics (ROC) Analyses.

### Likelihood Ratio Tests

Likelihood ratios and kappa values for detecting both depression and any mood disorder diagnosis across score ranges are reported in Table 5. Score ranges were computed by dividing the total distribution of scores on caregiver-report measures into quintiles, given the larger sample size for caregiver-report measures than for youth-report measures. For simplicity, the first two quintiles and the third and fourth were combined. For the CBCL and YSR, scores of 0– 2.9 were classified as “low risk”, 3–5.9 as “neutral risk”, and 6–10 as “high risk”. For the P-GBI and A-GBI, the low-, neutral-, and high-risk score ranges were 0–1.9, 2–5.9, and 6–15, respectively.

**Table 5.** Likelihood Ratios Associated with Test Scores on 5-Item Short Forms.

| Scale | Score Range | Risk | Any Mood Disorder |  | Depression |  |
| --- | --- | --- | --- | --- | --- | --- |
|  |  |  | Likelihood Ratio | Kappa | Likelihood Ratio | Kappa |
| CBCL | 0-2.9 | Low | .35 | -- | .45 | -- |
|  | 3-5.9 | Neutral | 1.12 | .28 | .93 | .11 |
|  | 6-10 | High | 2.65 | .20 | 2.05 | .21 |
| P-GBI | 0-1.9 | Low | .33 | -- | .44 | -- |
|  | 2-5.9 | Neutral | 1.16 | .35 | 1.05 | .14 |
|  | 6-15 | High | 4.13 | .27 | 1.99 | .20 |
| YSR | 0-2.9 | Low | .63 | -- | .69 | --- |
|  | 3-5.9 | Neutral | 1.05 | .20 | 1.04 | .13 |
|  | 6-10 | High | 3.33 | .15 | 1.92 | .14 |
| A-GBI | 0-1.9 | Low | .50 | -- | .50 | -- |
|  | 2-5.9 | Neutral | 1.02 | .20 | 1.09 | .14 |
|  | 6-15 | High | 1.72 | .16 | 1.42 | .13 |

*Note.* CBCL=Child Behavior Checklist; YSR=Youth Self-Report; P-GBI=Parent General Behavior Inventory; A-GBI=Adolescent General Behavior Inventory.

Likelihood ratios generally increased across score categories for each of the 5-item short forms. Low score ranges were associated with likelihood ratios below 1, indicating a reduced likelihood of either any mood disorder or a depression diagnosis. For the CBCL-5, P-GBI-5, and A-GBI-5, low scores decreased the likelihood of both any mood disorder and depression diagnoses by a factor of approximately 2 to 3, with likelihood ratios ranging from 0.33 to 0.50.

Likelihood ratios for the low score group as measured by the YSR-5 were closer to 1 but still substantially lower (any mood disorder LR=0.63, depression LR=0.69). Conversely, high score ranges were associated with likelihood ratios above 1, though the magnitude of likelihood ratios for any mood disorder diagnosis (LR=1.72-4.13) was greater than those for depression diagnosis (LR=1.42-2.05) in all cases.

Kappa values for each 5-item scale were calculated using binary classifications, with the low-scoring group as the referent. They indicated slight to fair agreement with diagnostic status, using the cutpoints suggested by Landis and Koch (1977). Kappa values were generally higher for any mood disorder diagnosis than for a depression diagnosis. Among the neutral-scoring groups, fair agreement with any mood disorder diagnosis was observed across all scales, with kappa values ranging from 0.20 to 0.35. Among the high-scoring groups, fair agreement with both any mood disorder and depression diagnoses was observed for the CBCL-5 and P-GBI-5, with kappa values ranging from 0.20 to 0.27.

## Discussion

In summary, we found evidence that short forms carved to retain coverage of five core symptoms of depression by matching VQIDS-5 items may provide informative and useful measures for shorter risk assessments. Evaluating short forms that use the VQIDS-5 as a template prioritizes content coverage over internal consistency. By testing multiple short forms carved from longer measures, we evaluated the robustness of using these five symptoms. Second, these short forms create linking item sets, in which the five items establish a crosswalk between datasets that have used different measures (Hussong et al., 2013). Lastly, these short forms offer an opportunity for settings that have invested in a tool (e.g., ASEBA, GBI) to switch to shorter versions, increasing uptake and reducing burden, while also avoiding the adoption friction of switching to an unfamiliar alternative (Beidas et al., 2025).

In the present study, all short forms reduced the total item count by more than 80%, indicating that they could substantially reduce the assessment burden for clinicians, youth, and caregivers. Further, all short forms correlated highly with the full-length measures, indicating that they capture information similar to that of the long forms. When comparing the psychometric properties of the short forms to those of the full forms, the short forms demonstrated similar or higher average-item correlations, which provide a purer measure of internal consistency because they are not influenced by scale length (Streiner et al., 2015).

Support for the criterion validity of the short forms was strong. The CBCL-5, YSR-5, P-GBI-5, and A-GBI-5 did not differ from their respective full-length measures in their correlations with various criteria, including psychiatric diagnoses and clinical interviews of depression and other mood disorders, to a practically meaningful extent. The discrepancies in validity coefficients were negligible or small based on Cohen’s *q*-statistic benchmarks (Cohen, 1988; Youngstrom et al., 2019).

When evaluating the diagnostic utility of the short forms, AUC values were generally not meaningfully lower than those of their corresponding long forms. When predicting any mood disorder or an MDD diagnosis, the short and full-length versions of the YSR and A-GBI did not differ significantly in diagnostic performance. The P-GBI-5’s discriminative validity was comparable to that of the 10-item depression short forms evaluated by Youngstrom et al. (2020, 2021), who reported AUCs of .68-.84 for the academic sample and .63-.80 for the community sample. The largest AUC difference was observed when the P-GBI-5 predicted any mood disorder, a comparison that pitted a 5-item versus a 46-item scale and included cases with bipolar disorders in the “any mood” reference group. The 46 GBI items include many designed to assess “biphasic” or mixed mood presentations, enhancing their ability to detect bipolar spectrum disorders. Additionally, diagnostic utility appears to be greater for caregiver-report measures than for youth-report measures, aligning with previous work underscoring the importance of caregiver report in PBD diagnosis (Youngstrom et al., 2015b). Thus, although the 5-item versions of these scales did not perform worse than the corresponding full-length measures on many criteria, use-cases that prioritize diagnostic classification and accuracy might be better served by longer alternatives.

Calculating likelihood ratios further underscored findings on diagnostic utility. Diagnostic likelihood ratios (DLRs) for low-and high-scoring groups were generally larger in magnitude for any mood disorder diagnosis than for depression specifically, and for caregiver-report measures than for youth self-report measures. These findings align with prior literature highlighting the utility of the P-GBI in screening for mood disorder symptoms more broadly (rather than for MDD alone) and the value of caregiver-report measures in providing useful clinical insight into youth symptoms. Further, our sample included a high rate of youth with bipolar disorder, likely inflating false positives among youth categorized as depressed. These individuals may have increased the false-positive rate for depression diagnosis but not for any mood disorder diagnosis.

When evaluating likelihood ratios across severity categories, DLR values greater than 2 (or less than 0.5) are generally considered moderately helpful for identifying clinically relevant differences in symptomatology (Youngstrom et al., 2015a). Our results indicate that high scores on the CBCL-5, P-GBI-5, and YSR-5, and low scores on the CBCL-5, P-GBI-5, and A-GBI-5, are likely to be informative about mood disorder symptoms broadly. Additionally, our results support the clinical utility of high CBCL-5 scores and low CBCL-5 and P-GBI-5 scores for identifying meaningful differences in MDD symptoms specifically. Most broadly, these scales are likely to be helpful in clinical settings for informing diagnosis and monitoring symptom change, but they should not be used in isolation to establish mood disorder diagnoses (Youngstrom, 2026).

Taken together, our results suggest that the CBCL-5, YSR-5, P-GBI-5, and A-GBI-5 perform comparably to the full-length measures from which they were derived. In clinical settings, these brief scales may be most useful when administered in combination with other evidence-based assessments rather than used in isolation. For example, when a child presents in a clinic with concerns from a caregiver that the child may be experiencing depression or another mood disorder, the child is already at a baseline risk of 17-26% for depression and 8-10% for dysthymia (Rettew et al., 2009). If that child has a family history of mood disorders, the risk of developing a mood disorder doubles, so the initial risk of depression upon presentation is up to 52% (Vandeleur et al., 2012). Although this risk is high, it is not sufficient to provide a diagnosis of a specific mood disorder, and more information is needed before a clinician decides to administer longer assessment tools. One tool that may provide such quick information is the short forms developed in the present study, which significantly reduce assessment time, provide information similar to the full-length forms, and indicate whether a child or adolescent may be at greater risk for a mood disorder.

### Strengths, Limitations, and Future Directions

A major strength is the sample size and its diversity in race, treatment setting, and severity, which enhance generalizability. Furthermore, diagnoses were determined using the KSADS and a consensus review, thereby providing research-grade criteria for evaluating the diagnostic utility of these short forms. Several limitations warrant consideration. First, selecting items from the full-length measures was sometimes ambiguous, particularly when multiple YSR, P-GBI, or A-GBI items matched the same target symptom. In these cases, we selected the item with the strongest EFA loading. Second, because this was a secondary analysis, we evaluated short-form items embedded within the full measures; their performance could differ when administered independently. However, the specificity and detail of GBI items may limit context effects, suggesting that independently administered P-GBI-5 and A-GBI-5 scores would be similar (Freeman et al., 2012). Direct administration of the stand-alone short forms is nevertheless needed to confirm this assumption. Third, comparisons between short and full forms involved multiple tests, including Cohen’s *q* tests of differences in criterion correlations and DeLong tests of differences in diagnostic accuracy, increasing the risk of Type I error. We applied Bonferroni correction to the criterion-correlation analyses, but some observed differences may still reflect multiple testing rather than true performance differences. Fourth, classification accuracy may vary across settings with different case mixes, symptom severity, and mood-disorder prevalence (Youngstrom et al., 2006, 2015a). Performance therefore should be examined in inpatient and general-population samples, as well as in clinical samples in which other disorders, such as anxiety disorders, may produce overlapping mood symptoms (Kessler et al., 2012). Finally, nationally representative norms are unavailable for these short forms. Existing CBCL and YSR norms apply only to their official scales, and the GBI has not been normed in a nationally representative sample, limiting interpretation of scores in the present sample.

Given these limitations, our study suggests a few potential avenues for future research. First, progress monitoring is a central use case for brief scales in measurement-based care. Evaluating the treatment sensitivity of these 5-item short forms would be a valuable, logical next step. In addition, the hypothesis that the five symptoms provide sufficiently broad coverage of the depression construct could be tested by extracting alternative 5-item sets from the item pools of other depression scales to see whether they yield properties similar to those of the short forms developed here. This test would provide additional support for the validity of these five symptoms as core depressive symptoms. Because of copyright and commercial considerations, the 5-item forms extracted from the CBCL and YSR remain in the realm of research. In contrast, the GBI forms are part of a set of short variants that are being validated and translated across a variety of languages and settings.

## CRediT Authorship Contribution Statement

Allison M. Loomis: Conceptualization, Formal analysis, Writing – original draft.

Phoebe A. Rodda: Conceptualization, Formal analysis, Validation, Writing – original draft.

Jennifer K. Youngstrom: Investigation, Supervision, Writing – review & editing.

Robert L. Findling: Funding acquisition, Writing – review & editing.

A. John Rush: Writing – Conceptualization; Writing --review & editing.

Eric A. Youngstrom: Conceptualization, Funding acquisition, Project administration, Supervision, Writing-review & editing.

## Declaration of Generative AI and AI-Assisted Technologies in the Manuscript Preparation Process

During the preparation of this work, the authors used Claude Sonnet 5, ChatGPT-5.6 Luna, and Grammarlyduk to correct grammatical errors, improve conciseness, and identify areas that needed expansion or further explanation. After using these tools, the authors reviewed and edited the content as needed and take full responsibility for the content of the published article.

## Data Availability

All data produced in the present study are available upon reasonable request to the authors.

