## Supplemental Table 1 for "Evaluating 5-Item Short Forms of Youth Depression Measures"

**Supplementary Material**

**S1**. Contender Items and Factor Loadings for the P-GBI-5

| **Core Symptom** | **Item Number** | **Item** | **Factor Loading** |
| --- | --- | --- | --- |
| Low Mood | 03 | “Has your child become sad, depressed, or irritable for several days or more without really understanding why?” | .70 |
|  | 23 | “Have there been times of several days or more when your child was so sad that it was quite painful for him/her, or he/she felt that he/she couldn’t stand it?” | .76 |
|  | 34 | “Over the past year, have there been long periods in your child’s life when he/she felt sad, depressed, or irritable most of the time?” | .74 |
|  | **63** | **“Have there been periods lasting several days or more when your child was so down in the dumps that he/she thought he/she might never snap out of it?”** | **.77** |
|  | 72 | “Have there been periods of time when your child felt a persistent sense of gloom?” | .74 |
| Concentration Difficulties | **12** | **“Have there been times when your child’s memory or concentration seemed especially poor and he/she found it difficult, for example, to read or follow a TV program, even though he/she tried?”** | **.45** |
|  | 08 | “Have there been periods of several days or more when your child could not keep his/her attention on any one thing for more than a few seconds, and his/her mind jumped rapidly from one thought to another or to things around him/her?” | .32 |
|  | 01 | “Have there been periods in your child’s life over the past year when it was almost impossible to make small decisions even though this may not be generally true of him/her?” | .40 |
| Low Self-Worth | 32 | “Over the past year, have there been times when your child looked back over his/her life and could see only failures or hardships?” | .65 |
|  | 47 | “Have there been times when your child hated himself/herself or felt that he/she was stupid, ugly, unlovable, or useless?” | .63 |
|  | **56** | **“Have there been times of several days or more when your child really got down on himself/herself and felt worthless?”** | **.76** |
| Loss of Interest | **13** | **“Have there been times when your child lost almost all interest in the things that he/she usually likes to do (such as hobbies, school, work, entertainment)?”** | **.63** |
| Low Energy | 18 | “Have there been times of several days or more when your child was so tired and worn out that it was very difficult or even impossible to do his/her normal everyday activities (not including times of intense exercise, physical illness, or heavy work schedules)?” | .64 |
|  | **26** | **“Has your child had periods when he/she was so down that he/she found it hard to start talking or that talking took too much energy?”** | **.65** |
|  | 33 | “Has your child experienced times of several days or more when he/she felt as if he/she was moving in slow motion?” | .56 |
|  | 59 | “Have there been times when your child hated himself/herself or felt that he/she was stupid, ugly, unlovable, or useless?” | .63 |

*Note.* Items in bold had the strongest factor loadings within their symptom category and were therefore included in the GBI short forms (i.e., P-GBI-5 and A-GBI-5). Factor loadings were obtained using Principal Axis Factor Analysis with one factor.
